# What’s in a diagnosis? A genetic decomposition of major depression

**DOI:** 10.1101/2020.12.15.20247015

**Authors:** Bradley S Jermy, Kylie P Glanville, Jonathan RI Coleman, Cathryn M Lewis, Evangelos Vassos

**Affiliations:** Social, Genetic and Developmental Psychiatry Centre, Institute of Psychiatry, Psychology & Neuroscience, King’s College London, London, UK; NIHR Maudsley Biomedical Research Centre, South London and Maudsley NHS Trust, London, UK; Department of Medical & Molecular Genetics, Faculty of Life Sciences and Medicine, King’s College London, London, UK

## Abstract

Determining a diagnosis of major depressive disorder (MDD) is complex, involving consideration and rating of a variety of different components. These include number of symptoms over an agreed threshold, symptom duration, functional impairment, persistence of symptoms within an episode, and symptom recurrence. While these components are generally accepted amongst physicians, it is unknown whether they reflect partly distinct biology between phenotypes. The aim of this study was to investigate how the genetic aetiology varies in the presence of different MDD components.

Thirty-two depression phenotypes which systematically incorporate the MDD components were created using the mental health questionnaire data within the UK Biobank. SNP-based heritabilities and genetic correlations with three previously defined major depression phenotypes were calculated (broad depression, Psychiatric Genomics Consortium (PGC) defined depression and 23andMe, Inc. self-reported depression) and differences between estimates analysed.

All phenotypes were heritable (h^2^_SNP_ range: 0.102 – 0.162) and showed substantial genetic correlations with other major depression phenotypes (Rg range: 0.651 – 0.894 (PGC); 0.652 – 0.837 (23andMe); 0.699 – 0.900 (broad depression)). The requirement for 5 or more symptoms and for a long episode duration had the strongest effect on SNP-based heritability, in the positive and negative direction respectively (1.4% average increase; 2.7% average decrease). No significant differences were noted between genetic correlations.

While there is some variation, the two cardinal symptoms, depressed mood and anhedonia, largely reflect the genetic aetiology of phenotypes incorporating more MDD components. These components may appropriately index for severity, however, the genetic component between phenotypes incorporating none and all components is comparable.

## Introduction

Major Depressive Disorder (MDD) is a common mental health condition and a leading cause of disability (1). Diagnosing MDD is a complicated process as one relies on symptoms commonly observed in the population, the presentation of which should exceed a normal reaction to the patient’s current environment (2,3).

Qualitative studies have investigated which factors primary care physicians consider when evaluating a diagnosis of MDD. Many consistent themes arise, suggesting the presence of biological symptoms, e.g. weight loss or gain, context of the depression, functional impairment, episode duration, and recurrence as key differentiators (4–7). Given physicians refer to guidelines to make a diagnosis, it is not surprising these findings largely mirror criterion in the Diagnostic and Statistical Manual 5 (DSM5) and International Classification of Diseases 11 (2,3). Different weightings of these components between physicians lead to varied opinions about both the diagnosis and severity of MDD. Indeed, the DSM-5 field trials show a low diagnostic inter-rater reliability between clinicians (kappa=0.28, 95% CI: 0.20 – 0.35) (8).

If these components do facilitate a valid conceptualisation of MDD, it may imply the existence of biological differences between MDD and a lenient depression phenotype which does not consider the components. This is important to determine as if such differences exist, it provides biological support to the validity of the current diagnostic criterion.

Recent advances in biobank level data and genetic research allow us to explore the biological basis of diverse MDD presentations. Major depression (MD) has a significant heritable component, with estimates in the range of 20 and 50% (9–12). It is a polygenic trait, meaning the genetic variance is explained partly by multiple common genetic variants of individually small effect in the population (13). Due to its genetic architecture, large sample sizes of MD cases and controls are required to identify these variants (14). As such, many genome-wide association studies (GWAS) have used a pragmatic ‘minimal phenotyping’ approach whereby cases of MD are identified according to a single, self-report, questionnaire response.

The MD GWAS meta-analysis by Wray et al., (15) which identified 44 significantly associated risk variants, included a phenotype from 23andMe which used six versions of a self-report question asking if the participant has ever been diagnosed with clinical depression (16). In addition, the Howard et al., (17) GWAS meta-analysis included a ‘broad depression’ phenotype derived in the UK Biobank through self-report responses to the question ‘have you ever seen a general practitioner/psychiatrist for nerves, anxiety or depression?’ (18). This approach has been justified through the high genetic correlations shown between these minimal phenotypes and the current gold standard MD phenotype of European ancestries; namely the Psychiatric Genomics Consortium (PGC) which contain a large number of cases identified according to a structured diagnostic interview. The genetic correlations are 0.85 (SE=0.03) and 0.87 (SE=0.04) for the 23andMe and broad depression phenotypes respectively (17).

These results may support the use of the two minimal phenotypes (23andMe and broad depression) as they show a significant proportion of the genetic variants associated with a stricter MD phenotype are shared. Alternatively, as the correlations with the gold standard PGC phenotypes are significantly different from unity, these minimal phenotypes may reveal different genetic loci, indicating partly distinct phenotypes from a biological perspective. Cai et al., (19) provide a comprehensive analysis of the minimal phenotyping approach in genetics. Using the UK Biobank, multiple minimal phenotypes are compared to a strict definition of MD, derived from responses to the Composite International Diagnostic Interview – Short Form (CIDI-SF) (20). Relative to the minimal phenotypes, the strict MD phenotypes show higher SNP-based heritability and genetic correlations between minimally- and strictly-defined MD phenotypes differ significantly from 1.

These results may be interpreted in two ways. Firstly, the minimal phenotypes may include additional information not relevant or specific to a diagnosis of depression, such as treatment seeking or anxiety. However, it may also be that minimal phenotypes captures milder cases that would not meet full diagnostic criteria, or have not been systematically assessed for components of MD. For example, episode recurrence may reflect a higher burden of, and potentially involve some different biological pathways to single episode presentation. In this scenario, focusing on recurrent depression would result in an increased SNP-based heritability and genetic correlations with gold standard MD phenotypes relative to single episode depression, as shown by Cai et al., (19). Interestingly, if this interpretation is true, the genetic aetiology of the MD phenotype may vary with recurrence, leading to the possibility of specific biological pathways influencing this component.

To our knowledge, no study has systematically investigated this interpretation. We used genetic analyses to determine whether different constellations of MD components reveal variation in the genetic aetiology. According to the diagnostic criteria for MD and the considerations from primary care providers shown in the literature, we break down MD into 5 key components: (1) the presence of five or more of the nine depressive symptoms listed in DSM-5, (2) functional impairment, (3) episode duration (4) the persistence of depression during the episode, and (5) recurrence. Note that for symptom number, the count was inclusive of the two cardinal symptoms, depressed mood and anhedonia, with at least one cardinal symptom having to be endorsed in all cases.

This study used mental health data from the UK Biobank to define thirty-two depression phenotypes which systematically incorporate the five components. Through assessing patterns in SNP-based heritability and genetic correlations between the thirty-two depression phenotypes and the current European ancestries gold standard PGC MDD cohort, we aim to explore how the genetic aetiology of a MD phenotype varies in the presence of the five components. As a secondary aim, we repeated the genetic correlation analysis with two minimal phenotypes (23andMe self-reported and broad depression) to determine if these definitions show differential patterns to the PGC cohort, consistent with minimal phenotypes accounting for the MD components to differing degrees.

## Methods

### Data

The UK Biobank, a health study of 502,655 individuals, was used for this study (21). We used responses to the CIDI-SF which formed part of the Mental Health Questionnaire (MHQ) to define our phenotypes (20,22). This voluntary web-based questionnaire was completed by 157,366 UK Biobank participants aged between 45 and 82 when completing the questionnaire.

### Characterisation of the Phenotypes

The CIDI-SF contains questions relating to an individual’s worst episode of depression during their lifetime (20). Five components for MD were defined from CIDI-SF questions, corresponding to: episode recurrence (2 or more depressive episodes in lifetime), the presence of five or more depressive symptoms, a long episode duration (episode > 6 months), the presence of functional impairment (affected life/activities either ‘somewhat’ or ‘a lot’) and the persistence of the depressive symptoms during the episode (felt depressed ‘almost every day’ or ‘every day’). For brevity, these components will be referred to as recurrence, symptoms, duration, impairment, and persistence respectively. For each component, we derived a binary variable indicating if the individual endorsed this particular aspect of depression. For more detail as to how these binary variables were defined please see Supplementary Table 1.

A base phenotype was first created, identifying individuals who endorsed at least one of the cardinal symptoms of depression (depressed mood, anhedonia). In these individuals, presence or absence of each of the 5 binary components were then determined. Each individual could endorse between 0 and 5 of the components, resulting in a total of 32 different phenotypes shown by calculating 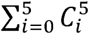 (Figure 1). The naming convention for each phenotype throughout the rest of the paper relates to which components are endorsed to be designated case status (Supplementary Table 2). For example, the phenotype ‘Cardinal+Recurrence+Impairment’ reflects all individuals who endorsed at least one cardinal symptom, reports more than one major depressive episode and was at least somewhat functionally impaired as a result of their worst episode. We use the term ‘enrichment’ to refer to the number of phenotypic factors endorsed, in addition to the cardinal symptoms (between 0 and 5).

**Figure 1:**
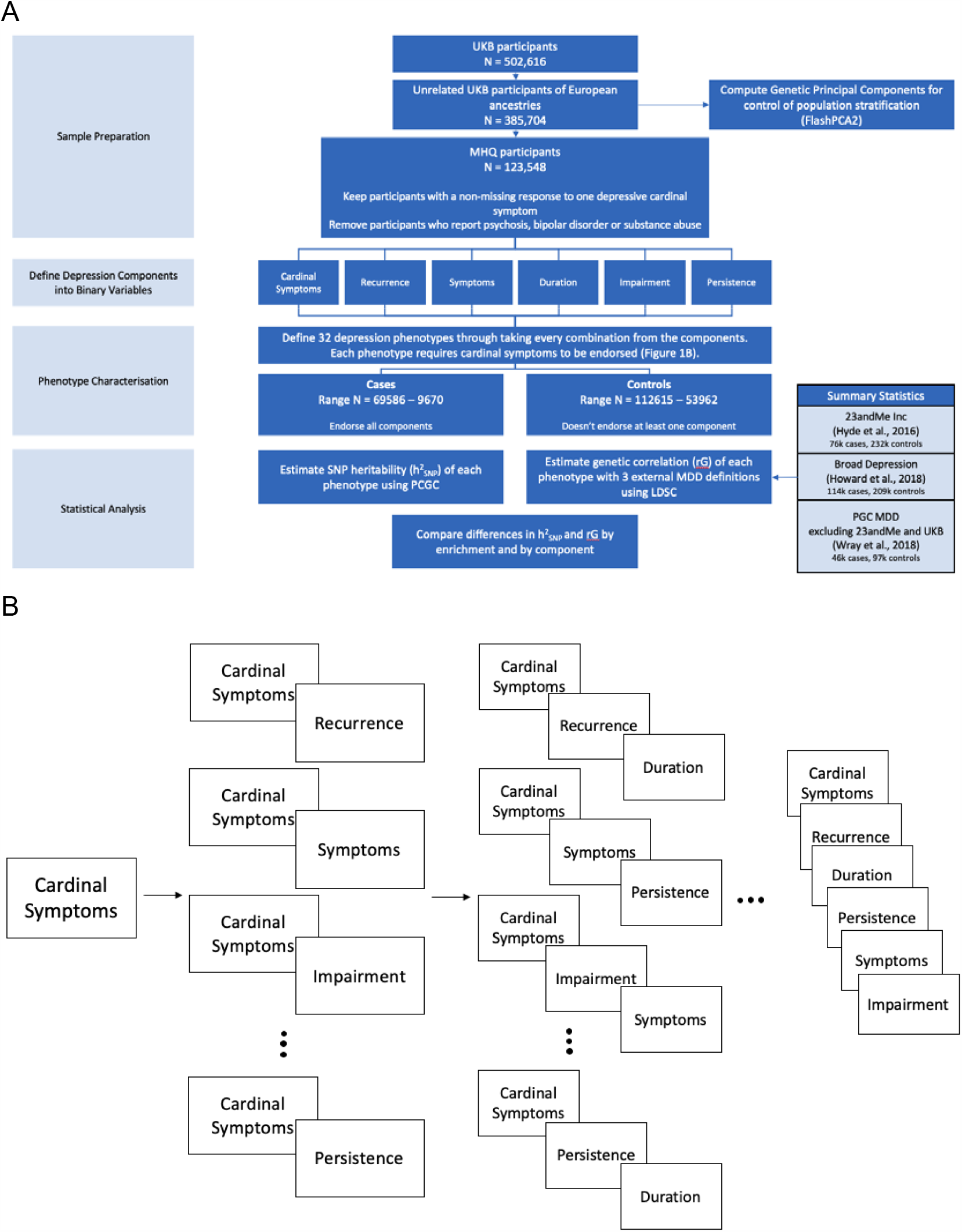
Workflow of the study design. **A.) Flow chart of key methodological steps.** Abbreviations: UKB – UK Biobank, PCGC – Phenotype Correlation Genotype Correlation, LDSC – Linkage Disequilibrium Score Regression, PGC – Psychiatric Genomics Consortium, MDD – Major Depressive Disorder **B.) Characterisation of the 32 phenotypes**. Figure 1B provides a graphical image of each phenotypes composition. Each time a component is added an additional phenotype is defined. Taking all possible combinations from each addition creates a possible 32 distinct phenotypes. The ellipses have been included in the graph to represent the additional phenotypes not included within the figure.

Participants were designated as a control if they did not endorse a single item required for the phenotype. For example, in the instance cardinal symptoms, recurrence and impairment are required for case status, as long as any of the three components are not endorsed, the participant would be designated as a control. Therefore, as a depression phenotype includes more components, the number of controls also increases due to more participants not endorsing a particular component (Supplementary Table 3). For an evaluation of this approach to defining controls and its influence on the results, refer to the Supplementary Information. Controls were not ‘double-screened’ for the presence of any other psychiatric disorders, including MD, as to avoid upwardly biasing the SNP-based heritability and genetic correlation estimates (23–25). However, participants were excluded independently of case/control status if they were identified as a possible case for schizophrenia, bipolar disorder or substance abuse (N_excluded_ = 3,032). This was determined through an individual self-reporting either the disorder or a relevant medication (Supplementary Table 4).

### Genetic Data – Quality control, SNP-based heritability and genetic correlations

#### Quality Control

Participants in the final sample were unrelated and of European ancestries which were identified using a previously described analytical pipeline (Supplementary Methods) (21,26,27). A total of 560,173 genotyped and 9,940,918 imputed SNPs remained after QC. Genotyped SNPs were used to estimate heritabilities and imputed SNPs were used to compute genetic correlations.

#### SNP-based Heritability

Phenotype Correlation-Genetic Correlation (PCGC; https://github.com/omerwe/S-PCGC) was used to estimate the SNP-based heritability of the 32 depression phenotypes. It is robust to sample ascertainment and non-normal effects of covariates in case-control studies (28,29). PCGC estimates SNP-based heritability by regressing phenotypic correlations on the genetic correlations between each pair of individuals while accounting for the effects of specified covariates. To convert to the liability scale, population prevalence was assumed to equal the sample prevalence prior to the application of any exclusion criteria for each of the 32 phenotypes (Supplementary Table 3). As recommended, the Major Histocompatibility Complex (MHC) region was removed (chromosome 6; 28,866,528 bp – 33,775,446 bp) reducing the total number of SNPs used to estimate the SNP-based heritability to 554,059 (30). The first 6 genetic principal components, genotyping batch and assessment centre were included as covariates for all phenotypes.

#### Genetic Correlation

Genetic correlations were computed using linkage disequilibrium score regression (LDSC) (31). LDSC was chosen for this analysis as the summary statistics necessary for PCGC estimation were not available for the three external MD phenotypes and we did not have access to the individual level data to compute these. Correlations were estimated for each of the 32 phenotypes with three depression phenotypes (UK Biobank broad depression (18), 23andMe self-reported depression (16) and PGC defined depression (15)). Summary statistics from Howard et al., (2018) (N_cases_=113,769, N_controls_=208,811), Hyde et al., (2016) (N_cases_=75,607, N_controls_=231,747) and Wray et al., (2018) with 23andMe and UK Biobank samples removed (N_cases_=45,396, N_controls_=97,250) were used to calculate the genetic correlations with the depression phenotypes. The 32 depression phenotypes were residualised by the first 6 genetic principal components, genotyping batch and assessment centre, then a GWAS performed using PLINK 2.0 (<u>cog-genomics.org/plink/2.0</u>,32,33) to obtain summary statistics for each phenotype. Pre-computed linkage disequilibrium (LD) scores, HapMap3 SNPs and the default settings of LDSC were used to calculate the genetic correlations for all phenotypes.

### Statistical Analysis

#### How does Enriching the Major Depression Phenotype Impact SNP-based Heritability and Genetic Correlation?

We investigated the trend in SNP-based heritability and genetic correlations with enrichment of the MD phenotype, by requiring endorsement of additional MD components into the phenotype. Using the phenotype requiring only cardinal symptoms as a reference, we tested for significant differences in SNP-based heritability and genetic correlation estimates for the remaining 31 MD phenotypes. This was performed using a previously described block jackknife methodology with 200 blocks (Supplementary Methods) (34,35). Differences from phenotypes with the same level of enrichment (i.e. endorsing the same number of components) were averaged by taking the inverse-variance weighted mean of the SNP-based heritability (or genetic correlations) (Supplementary Methods). For example, all phenotypes requiring cardinal symptoms and one other component (recurrence, symptoms etc.) would have the differences in SNP-based heritability and genetic correlation averaged to estimate the mean difference produced at that level of enrichment.

#### Component Importance

To investigate the relative effect of each component in driving any pattern in the SNP-based heritability and genetic correlation estimates, an analysis of differences by component was performed. For this test, differences in estimates are calculated using the same block jackknife approach, however, the phenotype requiring the endorsement of cardinal symptoms is no longer the reference. Instead, differences are calculated between all combinations of phenotypes which differ by only one component. For example, the two phenotypes; ‘cardinal symptoms + recurrence’ and ‘cardinal symptoms + recurrence + symptoms’ would be compared as the phenotypes are identical other than for the symptoms component. Of the two phenotypes being compared, the phenotype which includes the fewest components is set as the reference and any difference in estimates is attributed to the component that differs between the two phenotypes (symptoms, in the example above). All estimates were grouped according to the level of enrichment in the non-reference phenotype. Following this, each estimate is further grouped according to the component responsible for driving the difference and the inverse-variance weighted mean and standard error is computed.

## Results

### Phenotypes

From the 123,548 unrelated UKB participants of European ancestries who provided at least one non-missing response to the cardinal symptom items within the MHQ, 65,586 endorsed at least one cardinal symptom, and 9,670 of these endorsed all five components. Final sample sizes for all phenotypes are shown in Supplementary Table 3.

### SNP-based Heritability

SNP-based heritability estimates for the 32 phenotypes ranged from 0.102 (SE=0.015, Phenotype = Cardinal+Impairment+Persistence+Duration to 0.162 (SE=0.014, Phenotype=Cardinal +Symptoms) (Figure 2a). All estimates were significantly different from 0 following Bonferroni correction for multiple testing (a_Bonferroni_ < 0.0016 (0.05/32 phenotypes)) (Supplementary Table 5).

**Figure 2:**
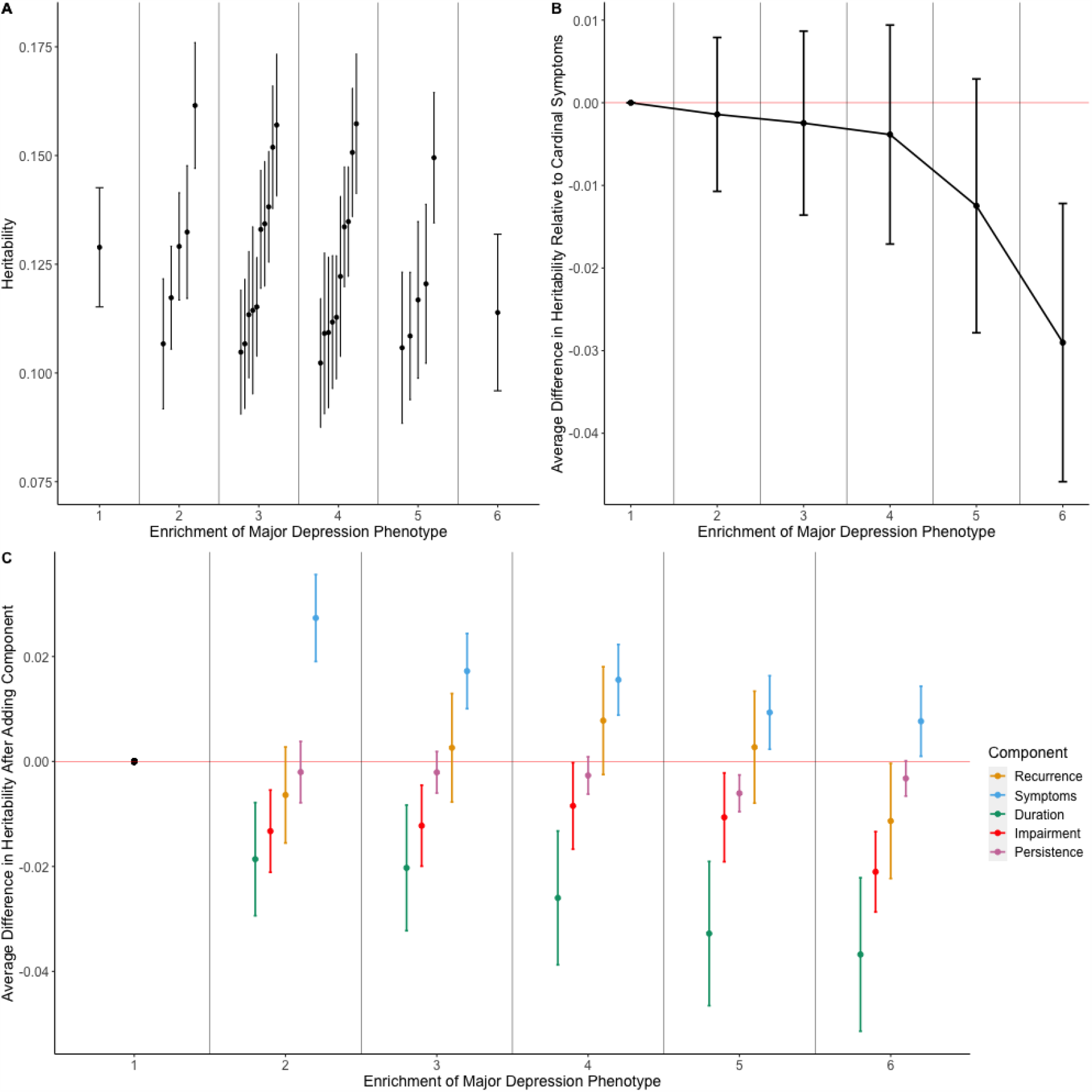
Analyses of SNP-based heritability of major depression phenotypes. **A.) SNP-based heritability estimates on the liability scale for each phenotype grouped by phenotype enrichment**. Phenotype enrichment is defined as the number of components considered to define case status. **B.) Trend in SNP-based heritability estimates by phenotype enrichment**. Each point estimate represents the average difference in SNP-based heritability relative to the phenotype which requires either of the cardinal symptoms to be endorsed. The cardinal symptom phenotype is the reference under the phenotype enrichment level 1. Error bars represent standard errors of the difference in SNP-based heritability estimates and averages were taken as the inverse-variance weighted mean of the enrichment group. **C.) Trend in SNP-based heritability estimates by component**. Each point estimate represents the average difference in SNP-based heritability induced from the addition of the component. Estimates are further grouped by level of phenotype enrichment. The point estimate with a phenotype enrichment of 1 is the cardinal symptoms phenotype and as such does not show any change due to the presence of no additional components. It is included for completeness. Errors bars represent standard errors of the difference in SNP-based heritability estimates and averages were taken as the inverse-variance weighted mean from all component comparisons within the enrichment group.

#### Trend with Phenotypic Enrichment

To understand the effect of enriching the MD phenotype, differences between the SNP-based heritability estimates relative to the phenotype of only cardinal symptoms were computed and averaged by the phenotypic enrichment, i.e. the number of components. As the phenotypes become more enriched, the average SNP-based heritability of the phenotype decreases (Figure 2b). However, taking each level of enrichment in turn, none of the SNP-based heritabilities were significantly different from the SNP-based heritability of the phenotype with only cardinal symptoms (p > 0.05) (Supplementary Table 6).

#### Importance of each component

Analysis by enrichment averages all phenotypes by number of components, however, phenotypes within each enrichment group will vary in which MD components are required to be endorsed. As such, the averaging removes any differential effect within the enrichment level, akin to treating all components as equivalent. To understand the differential impact of each component on SNP-based heritability, we first calculated the change in SNP-based heritability after adding the component and averaged the differences within each level of enrichment.

The requirement of five or more symptoms during the episode induced a significant increase in SNP-based heritability when the component was added to the cardinal symptoms only phenotype after correcting for multiple testing (α_Bonferroni_ < 0.002 (25 tests = 5 components over 5 levels of enrichment)) (Difference in SNP-based heritability = 0.027; SE=0.008; p-value=9.67×10^-4^). Inclusion or exclusion of all other components made no significant differences when grouped by enrichment (Figure 2c; Supplementary Table 7). The lack of significance limits comparisons across components, however, episode duration decreased SNP-based heritability to the greatest degree with an average decrease in SNP-based heritability across all levels of enrichment of 2.7%. It is, therefore, likely this component is contributing greatest to the decrease in SNP-based heritability with increasing phenotype enrichment. Conversely the presence of five symptoms increased SNP-based heritability to the greatest degree, with an average increase of 1.4% (Supplementary Table 7).

### Genetic Correlation

All genetic correlations were significantly different from 0 following Bonferroni correction for multiple testing (α_Bonferroni_ < 0.0016 (0.05/32 phenotypes)) (Supplementary Table 8). Genetic correlations ranged between 0.651-0.895 for PGC defined depression, 0.652-0.837 for 23andMe self-reported depression and 0.699-0.900 for broad depression (Figure 3a).

**Figure 3:**
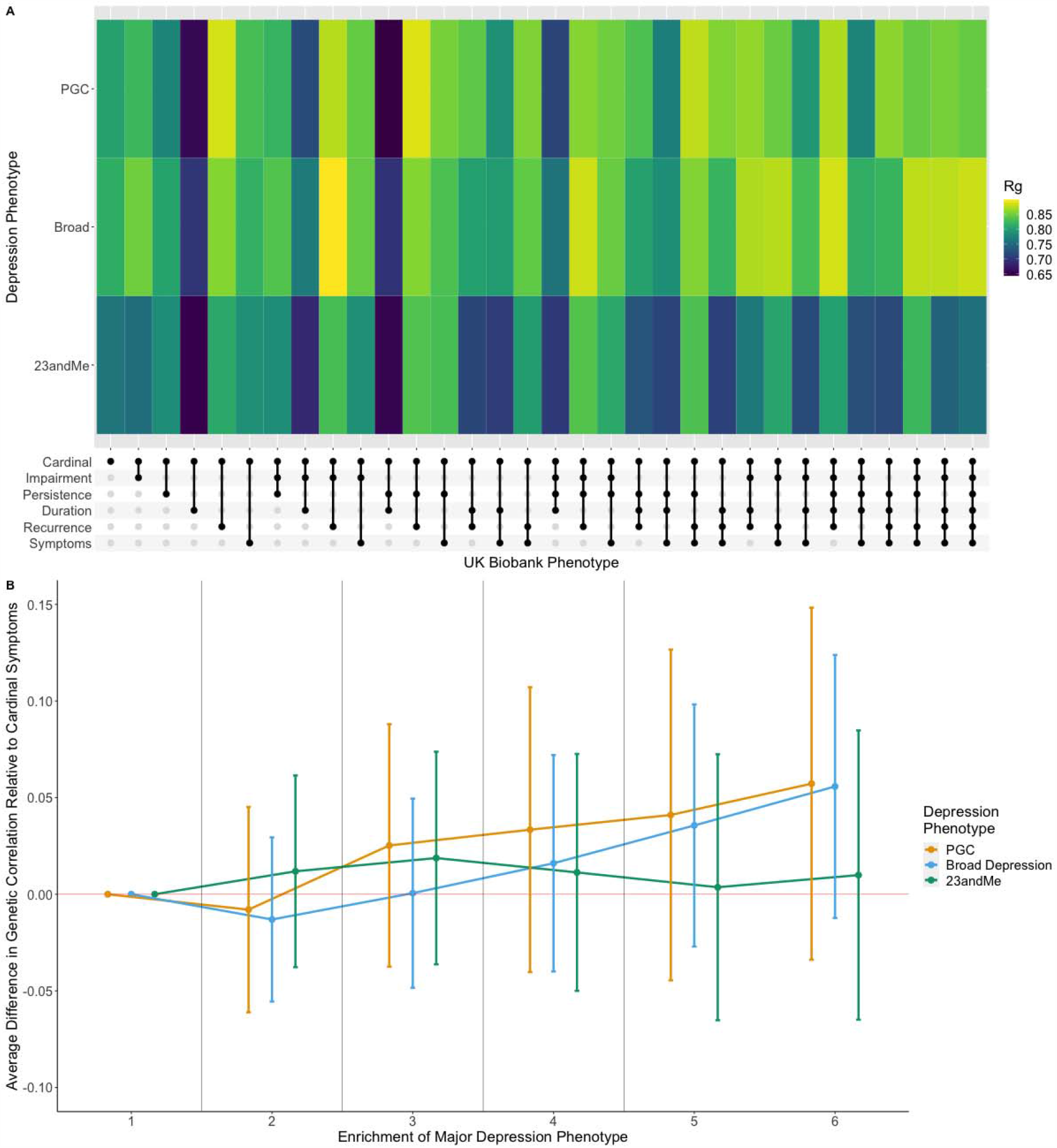
Analyses in genetic correlations with three previously defined major depression phenotypes. **A.) A heatmap of genetic correlation estimates for each phenotype**. Phenotypes we have defined are displayed on the x-axis. The three previously defined major depression phenotypes are displayed on the y-axis. Note: the legend shows the scale for correlation estimate comparisons is between 0.65 and 0.85. **B.) Trend in genetic correlation estimates with PGC defined major depression by phenotype enrichment**. Point estimates show the average difference in genetic correlation relative to the cardinal symptom only phenotype. This is shown as the reference point under the first level of phenotype enrichment. Error bars represent the standard errors of the difference.

#### Trend with Phenotypic Enrichment

Similar to the SNP-based heritability analyses, we analysed the effect of enriching the depression phenotype on the trend in genetic correlations with three external MD phenotypes. We compared all differences relative to the phenotype which required only cardinal symptoms to be endorsed. Differences in genetic correlation were not significant at any level of enrichment (p > 0.05) for broad depression, PGC or 23andMe defined depression. Both the broad depression and PGC defined depression showed an increase in genetic correlations estimates with enrichment of the depression phenotype, however, this is speculative and would require a study with greater power to show conclusively (Figure 3b; Supplementary Table 9). 23andMe self-reported depression did not show such an increase by phenotypic enrichment (Figure 3b; Supplementary Table 9)

#### Importance of each component

We analysed the change in genetic correlation induced by the addition of MD components for the three MD phenotypes. The addition of duration to the phenotype requiring only cardinal symptoms decreased the genetic correlation with all three of the MD phenotypes at a level of nominal significance (PGC: Δrg = -0.135; 23andMe: Δrg = -0.114; Broad depression: Δrg = -0.113; p < 0.05). Similarly, the addition of persistence to phenotypes that consisted of cardinal symptoms and one other component on average increased the genetic correlation with the 23andMe phenotype (23andMe: Δrg = 0.034, p < 0.05. However, none of these associations survive correction for multiple testing (α_Bonferroni_ < 0.002 (0.05/25)) (Figure 4; Supplementary Table 10).

**Figure 4.**
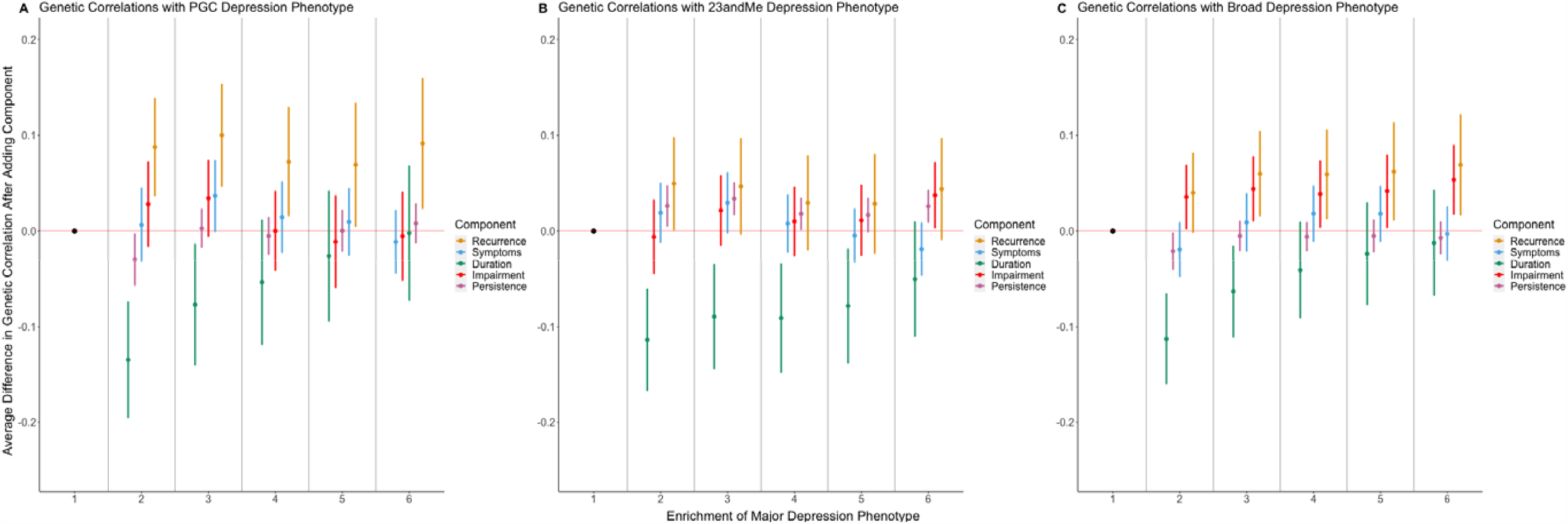
Trend in genetic correlation estimates by component. **A.) Trend using PGC defined major depression phenotype as the comparison for genetic correlation computation. B.) Trend using 23andMe defined major depression phenotype as the comparison for genetic correlation computation. C.) Trend using broad depression phenotype as the comparison for genetic correlation computation**. Each point estimate represents the average difference in genetic correlation induced from the addition of the component. Estimates are further grouped by level of phenotype enrichment. The point estimate with a phenotype enrichment of 1 is the cardinal symptoms only phenotype and as such does not show any change due to the presence of no additional components. It is included for completeness. Errors bars represent standard errors of the difference in genetic correlation estimates and averages were taken as the inverse-variance weighted mean from all component comparisons within the enrichment group.

Given this lack of association it is difficult to discern any real trend by component, however, incorporating recurrence into the depression phenotype resulted in the greatest average increase over all three of the depression phenotypes (average change: PGC = 8.8%; 23andMe = 4%; Broad depression = 6.4%). Conversely, incorporating episode duration into the depression phenotype consistently decreased the genetic correlation for all three depression phenotypes (average change: PGC = -5.1%; 23andMe = -8.5%; Broad depression = -4.4%).

## Discussion

In this study, we aimed to determine how the genetic aetiology varies between different conceptualisations of MD phenotypes. By defining five depression components in addition to the cardinal symptoms, we compared how the SNP-based heritability of the trait, and the genetic correlations with three previous depression studies varied with presence and absence of each component. Differences in SNP-based heritability would support a different risk factor profile, i.e. reduced effects of the environment in MD phenotypes with increased SNP-based heritability, whereas differences in genetic correlations would provide evidence for specific genetic variants contributing to the MD phenotype. Each piece of evidence could be used to support a partial biological distinction between phenotypes. We compared results across the 32 depression phenotypes defined, contrasting the effect of each component and the depth of enrichment, i.e. by how many components were endorsed.

Variability in SNP-based heritability across the phenotypes was low with a range of 5.9%. Relative to the cardinal symptom phenotype, the greatest increase in SNP-based heritability was 2.7%. Conversely, the greatest decrease was 3.9%. We caveat the latter with the fact that this difference is not statistically significant. This lack of variability suggests the genetic aetiology of the cardinal symptoms phenotype largely reflects that of other phenotypes which include more components of MD.

Interestingly, the phenotype with the highest SNP-based heritability was also parsimonious, requiring a cardinal symptom, and 5 or more symptoms during the episode. The increase in SNP-based heritability, from 12.9% for cardinal symptoms to 16.2% for cardinal symptoms and 5 or more symptoms was significant, which suggests the addition of 5 or more symptoms as a component leads to a more heritable phenotype, less influenced by environmental risk factors. The parsimonious nature of this phenotype, and the fact that richer phenotypes had lower SNP-based heritability, closer to that of the cardinal symptoms, is an important finding as it indicates much of the differentiable heritable signal for MD over and above the cardinal symptoms may be captured from a careful assessment of symptoms of depression. Thorp et al., (36) show SNP-based heritability varies between 6% and 9% for individual symptoms of MD. It is therefore plausible that further heterogeneity for SNP-based heritability within this component exists and that certain symptom profiles will be more heritable than others. Through simulating a phenotype according to the liability threshold model, Cai et al. (19) show that SNP-based heritability does not vary between milder and more extreme forms of the same phenotype, equivalent to lowering or increasing the threshold. In practice, accurately estimating SNP-based heritability depends on selecting an accurate population prevalence, akin to the threshold in the simulation. If this is mis-estimated, differences in estimates will arise artificially. The significance of our finding would be reduced if we have either under-estimated the cardinal symptom or over-estimated ‘Cardinal+Symptom’ population prevalence’s while holding the other phenotypes population prevalence constant.

While episode duration of greater than 6 months did not survive correction for multiple testing, the point estimates showed an effect of decreasing SNP-based heritability when incorporated into the phenotype. Indeed, a phenotype requiring the cardinal symptoms and a long episode duration is consistent with the depressive condition of dysthymia, which is considered a distinct diagnosis relative to more episodic depressive episodes in the DSM-5 (2). This finding is corroborated by a twin study which showed a negative relationship between monozygotic-dizygotic concordance and episode duration (37). A possible explanation for this is that episode duration is driven by environmental risk factors which increase the contribution from the environmental variance component. Indeed, socioeconomic status (SES) and marital status have been found to predict a longer duration (38), although other studies find no such association (39). Alternatively, duration of each episode may depend on response to treatment, which could be under different genetic and environmental influences.

Much of the literature exploring heritability by component has focused on recurrence, which has consistently been shown to increase the heritability of MD (9,11,12,19,40). Our results are therefore somewhat surprising in that we observe no increase in SNP-based heritability when incorporating recurrence, particularly given some of the previous literature used the same sample from the UK Biobank. The difference might be attributed to the definition of our control group which simply required a negative endorsement of a single component. This reduces the risk of a discontinuity when transforming to the liability scale which inflates SNP-based heritability estimates (23,25). However, the control cohort will also include participants considered ‘sub-threshold’ for each phenotype in which cardinal symptoms and possibly other MD components are endorsed. This increases the risk of bias due to misclassification (41) however we believe our definition finds a good balance between the two potential biases (see Supplementary Information for a more detailed discussion). The method used to assess lifetime MD (i.e. retrospective self-report vs prospective, structured, face to face interviews) may help to explain our results as differences in measurement error impact the results. While sample size is likely going to restrict an analysis using clinical interviews, future work attempting to replicate our findings using this method of assessment would shed light on the degree to which our results are driven by the retrospective, self-report nature of the assessment. Indeed, efforts to genotype the most severely depressed cases at scale will dramatically enhance our ability to unpick the heritability of MD (42).

To put our findings into further context, our results should not be used to draw conclusions on the relative power of each phenotype to detect genetic variants associated with MD. GWAS often use extreme cases and controls as a cost-effective way of improving power. In this scenario, if one is to assume MD along a continuum of severity and that these components are a proxy for severity, phenotypes which incorporate a greater number of components will likely enhance power for variant discovery (43). However, our study finds that the upper bound of prediction on extreme phenotypes in terms of the variance explained in the population is unlikely to be substantially greater than that for milder forms of depression. We also note the large number of tests we have performed in this exploratory analysis which will limit conclusions regarding significance. Future confirmatory studies with greater power, possibly targeting specific components to reduce multiple testing burden, may indeed reveal significant differences behind these phenotypes.

We have shown that the cardinal symptoms phenotype has comparable SNP-based heritability to the more enriched phenotypes of MD. A natural follow-up question is how specific each phenotype is relative to MD with respect to the genetic variants implicated. To test this, we calculated genetic correlations between each phenotype and three definitions of MD; PGC defined MD, 23andMe self-reported MD and broad depression.

The genetic correlations between the cardinal symptom only phenotype and the three depression phenotypes were high (PGC: rg=0.807, SE=0.054; 23andMe: rg=0.762, SE=0.041; Broad depression: rg=0.815, SE=0.032). Recurrence and duration were the two components to increase and decrease the genetic correlation point estimates, showing a consistent pattern across the three MD definitions. However, while there was variation around the correlation estimate given by the cardinal symptom phenotype (PGC: Range = -14.6% - 9.8%; 23andMe: Range= - 11.4% - 8.5%; Broad depression: Range = -12.1% - 7.8%), we found no evidence for a statistically significant increase or decrease either by phenotype enrichment or by component. As such, we cannot conclude that MD components change the specific set of associated genetic variants, which would support a partial distinction in biology between phenotypes. The relative increase that can be induced from the components is limited due to the high correlation between MD and the cardinal symptom only phenotype. As such, a ceiling effect is imposed which would require large sample sizes to detect significant differences for such correlations.

We had hypothesised that the two minimal phenotypes (the help-seeking broad depression in UK Biobank, and self-reported depression in 23andMe) are composed of a case sample with greater heterogeneity whereby the additional components have not been incorporated to the extent that they have with the gold standard PGC defined depression. Given the inclusion of potentially milder cases, we expected the minimal phenotypes to correlate with milder depression phenotypes to a greater extent relative to more enriched depression phenotypes. In contrast, we hypothesised PGC defined depression, being our gold standard, would show a positive trend between phenotypic enrichment and genetic correlation. However, no conclusive trend could be found by phenotype enrichment in any case. Interestingly, of the three depression genetic studies, broad depression showed the strongest trend with phenotype enrichment. As before, we caveat these trends with the note that no differences in genetic correlation were significant.

A key challenge in the study of the genetics of MD is to attain valid, comparable cohorts with genetic correlations that are not significantly different from unity. We hypothesised selection of cases according to a set of components may be important to produce homogenous cohorts. While the enrichment of the phenotypes with the components do seem to increase genetic correlations with previous MD definitions, there are additional avenues in which heterogeneity can manifest. We speculate that a consideration of the context behind an individual’s depression may be another component that would enhance the genetic correlations between cohorts (44). For example, grouping participants who present with a depressive episode following diagnosis of a chronic medical condition or following a stressful childbirth could reduce heterogeneity. This ‘splitting’ approach would be ideal in a world with an infinite population where access to all information regarding the depressive episode was available. However, a pragmatic approach must be employed as large samples are required to unpick the genetic aetiology of MD which may not be possible if we are to group cases by too many variables. Finding the sweet spot of this pragmatism is something that must be agreed between researchers and clinicians.

## Limitations

The results from this study should be evaluated in the context of the following limitations. To accurately estimate and compare heritabilities, the prevalence of the phenotype within the population must be estimated accurately. No previous literature exists for most of our phenotypes, so an assumption was made that UK Biobank represents a random subset of the population. This assumption is strong given participants of the UK Biobank have been shown to have a higher socio-economic status and lower mortality rates than the rest of the UK (45). The subset of participants who responded to the MHQ also have a lower rate of mental health related hospital diagnoses (46). Future consideration towards developing a representative dataset free from selection bias would help improve the validity of the prevalence’s used in this study.

All components used to define the phenotypes were dependent on the cardinal symptoms being endorsed as the questions relating to components such as recurrence were only asked in this instance. Understanding impairment distinct from the cardinal symptoms would be of interest however the nature of these questions would likely need to change as currently the impairment is attached to the depressive episode.

We assume the difference in SNP-based heritability and genetic correlation estimates is attributable to the component that has been changed between the two phenotypes. It is likely in practice that this component covaries with risk factors for depression and even other components such as depression. For example, should you endorse the five symptoms of depression you are also more likely to endorse recurrence. This limitation may be unpicked through investigating the set of participants who endorse one component but not the other, i.e. those that endorse recurrence but not the five symptoms, however, much greater sample sizes are required for such an analysis and the translational interpretation is less clear.

With respect to the genetic correlation analysis, we considered the PGC defined phenotype to be the gold standard for comparison against minimal phenotypes. It is indeed the case that this phenotype is the most stringently assessed for individuals of European ancestries, however, given MD’s inherent heterogeneity, it is unlikely all cases within this phenotype are recurrent or have had episodes of long duration. An equivalent external phenotype in which all components were known to be endorsed for all cases would be able to show more conclusively if the incorporation of the component provides more genetically comparable phenotypes. However, this is not how MD is currently defined in the diagnostic criterion so while more severe MD phenotypes (42,47) may behave as a better positive control for this study, it would only reflect a small subset of the total MD cases in the population.

## Conclusion

In this study we show that the key component in a MD phenotype from a genetic perspective are the two cardinal symptoms. We find evidence that out of the additional criteria typically used to establish diagnosis or severity of depression, incorporating five or more symptoms into the phenotype produces a significant increase in SNP-based heritability. While these components may be used to reduce misclassification between controls and cases and enhance power in GWAS, they do not appear key to identifying any distinct genetic aetiology of MD.

## Data Availability

The full GWAS summary statistics for the 23andMe discovery data set will be made available through 23andMe to qualified researchers under an agreement with 23andMe that protects the privacy of the 23andMe participants. Please visit https://research.23andme.com/collaborate/#dataset-access for more information and to for more information and to apply to access the data. All other GWAS summary statistics are publicly available from the following websites https://www.med.unc.edu/pgc/download-results/ (PGC summary statistics) and https://datashare.is.ed.ac.uk/handle/10283/3083 (broad depression summary statistics).  

https://www.med.unc.edu/pgc/download-results/

https://datashare.is.ed.ac.uk/handle/10283/3083

## Acknowledgements

CML is funded by the Medical Research Council (N015746/1). This study represents independent research funded by the National Institute for Health Research (NIHR) Biomedical Research Centre at South London and Maudsley NHS Foundation Trust and King’s College London. The views expressed are those of the author(s) and not necessarily those of the NHS, the NIHR or the Department of Health and Social Care.

We thank participants and scientists involved in making the UK Biobank resource available (http://www.ukbiobank.ac.uk/). UK Biobank data used in this study were obtained under approved application 18177. We thank the research participants and employees of 23andMe for making this work possible.

