## Supplementary Note for "What’s in a diagnosis? A genetic decomposition of major depression"

**Supplementary Methods**

Genotyping quality control procedures

Testing for significant differences in heritability and genetic correlations

Computing average differences in heritability and genetic correlations across phenotypes

Choosing unscreened controls

**Supplementary Figure 1: Heritability estimates using linkage disequilibrium score regression varied by how controls are defined and the method of converting to the liability scale.**

**Supplementary References**

**Supplementary Methods**

**Genotyping Quality Control Procedures**

An initial round of quality control (QC) was conducted centrally by the UK Biobank and is described in Bycroft et al., (1). Imputation was also performed centrally using two reference panels; the Haplotype Reference Consortium and UK10K (2,3). The sample was restricted to unrelated participants of European ancestries. Ancestry was determined via the largest cluster from 4 means clustering on the first two principal components generated from the genotype data centrally by the UK Biobank (4). Unrelatedness was set such that no two participants had a kinship coefficient r < 0.44 computed through KING with a greedy algorithm specified to retain the greatest sample from the exclusions (5).

Single Nucleotide Polymorphisms (SNPs) were restricted to have minor allele frequency > 0.01 and be in approximate Hardy-Weinberg equilibrium (test threshold: p < 10^-8^). For the imputed genetic data, only SNPs imputed with high confidence were retained such that INFO > 0.4. For participants, genetic sex, identified via X-chromosome homozygosity (FX) (FX > 0.9 for males, FX < 0.5 for females), was checked against self-reported sex and a < 2% overall SNP missingness threshold was imposed.

After QC in the full UK Biobank sample, 560,173 directly genotyped SNPs and 385,704 participants of European ancestries remained. This sample was used to calculate genetic principal components which were used to control for residual population stratification within European ancestries by Flashpca2 (6).

**Testing for significant differences in heritability and genetic correlations**

This methodology has been previously described by Hubel et al., (7). All notation is consistent with this publication for ease of understanding. To test for significant differences between two heritability and genetic correlation estimates, the original estimates to be compared are calculated; hereon defined as the ‘global estimate’.

A jackknife approach was then employed whereby a portion of the genome is masked proportional to the number of genotype blocks specified and the relevant estimate for the two phenotypes was re-calculated. We defined the number of blocks as 200, therefore, 200 different estimates were calculated from the jackknife. We denote the vector form (200 dimensions) of these estimates as R(A) and R(B) for estimates A and B respectively.

The difference between the two global estimates was calculated which will be a scalar value; denoted here as d(A,B). The difference between the two vectors R(A) and R(B) was denoted D(A,B). Jackknife pseudovalues were computed using d(A,B) and D(A,B) using the following formula:

P_i_(A,B)=n*d(A,B) - (n-1)*D_i_(A,B)

Where P_i_(A,B) represents the ith pseudovalue. In our instance we will calculate 200 pseudovalues given the dimensionality of the D(A,B) vector.

The mean and variances of the pseudovalues are respectively:

$$m(A,B)=\frac{1}{n}\sum_{i=1}^{n} P_{i}(A,B)$$

$$v(A,B)=\frac{1}{n-1}\sum_{i=1}^{n} {(P_{i}(A,B)-m(A,B))}^{2}$$

The mean and variance of the pseudovalues were used to derive a z-statistic and corresponding p-value to test the null hypothesis that there is no difference between the two estimates. The z-statistic is computed generally via the following formula:

$$z(A,B)=\frac{m(A,B)-\theta_{0}}{\sqrt{(1/n)\times v(A,B)}}$$

Where $\theta_{0}$ is the estimate you are comparing the observed difference to. In this instance, we were interested in testing whether the estimates deviate from 0 so specified the value as such.

**Computing average differences in heritability and genetic correlations across phenotypes**

To analyse the effect of enrichment and component, both SNP-based heritability and genetic correlations were averaged. To give an example, many differences in SNP-based heritability were conducted on phenotypes with one extra component (recurrence, symptoms etc.) relative to the cardinal symptoms. These were compared for significant differences versus the cardinal symptom phenotype. To get an understanding of what enriching the phenotype by one component does generally, we computed the average differences using an inverse variance weighted approach. The equations for the inverse variance weighted mean difference.

Inverse Variance Weighted Mean Difference = $\frac{\sum_{i=1}^{n} y_{i}/{\sigma_{i}}^{2}}{\sum_{i=0}^{n} 1/{\sigma_{i}}^{2}}$

where y_i_ represents the difference in estimates for each test and $\sigma_{i}$ represents the standard error of that estimate.

If we were to simply take the inverse variance weighted standard error, this would artificially reduce the standard error by placing more weight onto smaller standard errors within the equation. To avoid this, we instead calculated the total variance ${{(\sigma}_{i}}^{2})$ from the standard error of the estimates and multiplied this by a weighting factor as shown in the below equation (8).

Standard Error of Mean Difference = $\sqrt{\sum_{i=1}^{n} {\sigma_{i}}^{2}x \frac{(\sum_{i=1}^{n} {(1/{\sigma_{i}}^{2})}^{2})}{(\sum_{i=1}^{n} {(1/{\sigma_{i}}^{2}))}^{2}}}$

**Choosing between screened and unscreened controls**

Many genome-wide association studies sample from the extremes of the distribution in liability for cases and controls to improve power for variant detection (9). As shown by Yap et al., (10) this method can lead to upwards bias of heritability estimates as it will create a discontinuity in the liability distribution. This can be corrected through incorporating an additional population prevalence for the controls using the following equation (11).

$h_{l}^{2}=h_{o}^{2}\frac{1}{P(1-P)}\frac{1}{{(\frac{Z_{u}}{K_{u}}+ \frac{Z_{L}}{K_{L}})}^{2}}$	(equation 1)

where $h_{l}^{2}$ is the heritability on the liability scale, $h_{o}^{2}$ is the heritability on the observed scale, $P$ is the sample prevalence, and $K_{u}$and $Z_{u}$ represent the population prevalence of cases and the height of the standard normal curve at such a prevalence. Similarly, $K_{L}$and $Z_{L}$ represent the population prevalence of controls and the height of the standard normal curve at the specified prevalence.

We have chosen Phenotype-Correlation-Genotype-Correlation (PCGC) as our method for computing heritability as it has been designed specifically for binary phenotypes (12,13). The software, however, has built in the conversion to the liability scale and we therefore cannot impose the two-threshold equation highlighted above which would have been the ideal methodological decision. As such, a decision between screened and unscreened controls was taken according to an analysis using linkage disequilibrium score regression (LDSC).

In this analysis, screened controls are defined as controls that did not endorse either of the cardinal symptoms. As highlighted in the main manuscript, both cases and controls were screened for possible cases of bipolar disorder, schizophrenia and substance abuse. Unscreened controls do not have this restriction and therefore controls are defined as any individual that does not endorse a single diagnostic factor. In the scenario of the screened controls, increasing the number of diagnostic factors required for an individual to be defined as a case increases the degree to which the centre of the distribution in liability is removed which may inflate heritability estimates. The use of unscreened controls, however, does not remove the distribution in liability and is unlikely to suffer from inflated estimates due to this bias. We therefore hypothesised that converting to the liability scale using a single case prevalence in screened controls would lead to inflated heritability estimates and that correction with a control prevalence would alleviate this. We hypothesised that the unscreened controls would be much closer to these corrected estimates. In all scenarios cases were defined in the same way as the collection of individuals who endorse all necessary major depressive disorder components for the chosen phenotype (Supplementary Table 2).

To test this, we first derived heritability estimates on the observed scale using the unscreened controls and screened controls with LDSC. We then converted each heritability to the liability scale using the following equation which only requires the specification of the case population prevalence:

$h_{l}^{2}=h_{o}^{2}\frac{K(1-K)}{z^{2}}\frac{K(1-K)}{P(1-P)}$	(equation 2)

All variables within the equation have the same meaning as highlighted above however K and z now correspond to the case population prevalence and height at the standard normal curve in cases only.

We then converted the heritability estimates of the screened controls to the liability scale using equation 1 to understand which control definition produced estimates that were closest to this ideal methodological decision. The control prevalence was set as the proportion of participants within the Mental Health Questionnaire (MHQ) who endorsed at least one cardinal symptoms (43%). The results are shown in Supplementary Figure 1. As can be seen, prior to correcting for the prevalence of the controls, the use of screened controls provides inflated heritability estimates. Interestingly, the correction brings the estimates in line with that of unscreened controls. As such, a decision was taken to use unscreened controls for our analysis as this was deemed to be the methodological decision that minimised bias in our estimates.

We acknowledge that biases are likely to remain when using the unscreened controls. Incorporating the full sample is likely to increase the misclassification rate which has been shown to bias the heritability estimates downward (14). This is likely to cause bias in the analyses of differences in heritability across phenotypes if the misclassification rate changes between the 32 depression phenotypes. Further, we are restricted to using LDSC for our analysis as PCGC does not allow for different methods for converting to the liability scale. As such, we cannot exclude the possibility of different results using a different method.

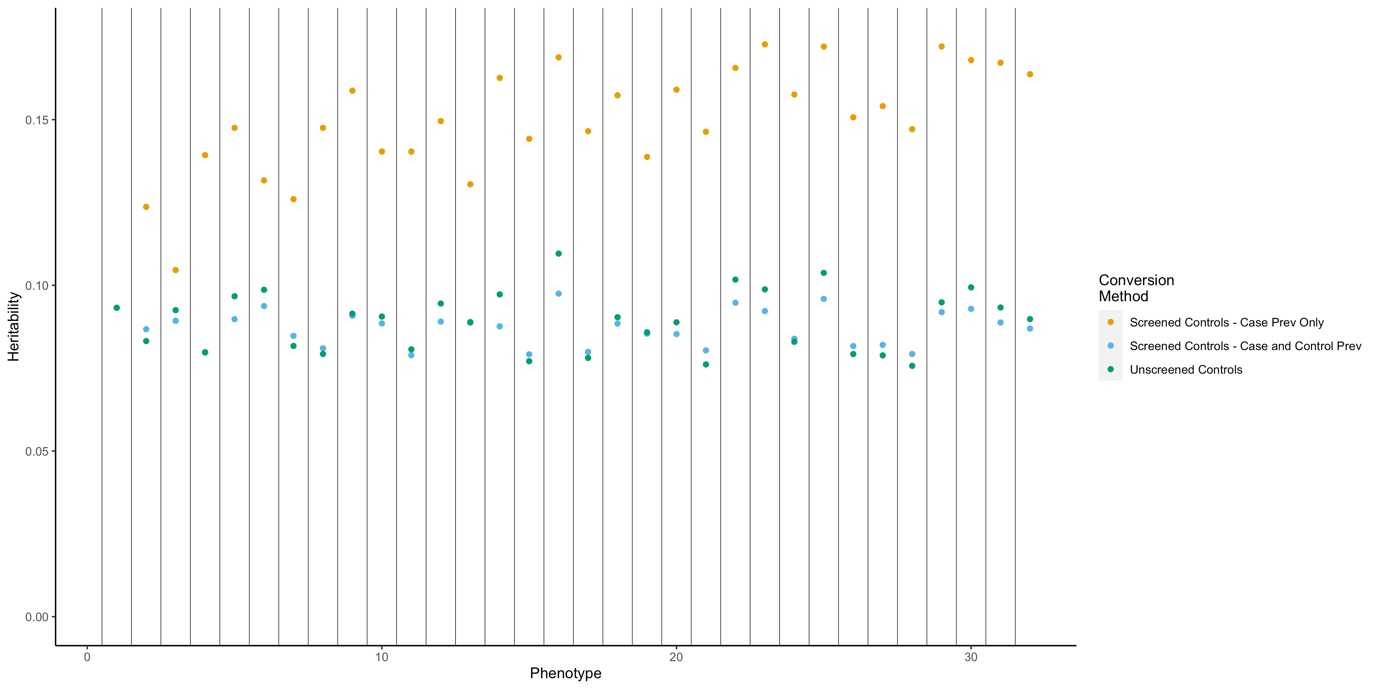

**Supplementary Figure 1: Heritability estimates using linkage disequilibrium score regression varied by the control definition and the method of converting to the liability scale.** Points in orange show heritability estimates when using screened controls and converting to the liability threshold with a single case population prevalence. Points in blue show heritability estimates using screened controls when a control population prevalence is included. Points in green show the heritability estimates derived using unscreened controls and a single case population prevalence. Phenotype number reflects the reference number found in supplementary table 2/
